# Health and Wellbeing Cohort Study of Serving and Ex-Serving UK Armed Forces Personnel: Phase 4 Protocol

**DOI:** 10.1101/2023.08.17.23294221

**Authors:** Marie-Louise Sharp, Margaret Jones, Ray Leal, Lisa Hull, Sofia Franchini, Niamh Molloy, Howard Burdett, Amos Simms, Steven Parkes, Daniel Leightley, Neil Greenberg, Dominic Murphy, Deirdre MacManus, Simon Wessely, Sharon A.M. Stevelink, Nicola T Fear

**Affiliations:** King’s Centre for Military Health Research, Department of Psychological Medicine, King’s College London, Weston Education Centre, 10 Cutcombe Road, London, UK; King’s Centre for Military Health Research, Department of Psychological Medicine, King’s College London, London, UK; King’s Centre for Military Health Research, Department of Psychological Medicine, King’s College London, London; Academic Department of Military Mental Health, Ministry of Defence, UK; Combat Stress, Research Department, Tyrwhitt House, Leatherhead, Surrey, UK; Department of Forensic and Neurodevelopmental Science, King’s College London, London; Academic Department of Military Mental Health and King’s Centre for Military Health Research, Department of Psychological Medicine, King’s College London, London, UK

## Abstract

**Introduction:** This is the fourth phase of a longitudinal cohort study (2022-2023) to investigate the health and wellbeing of UK serving (Regulars and Reservists) and ex-serving personnel (veterans) who served during the era of the Iraq and Afghanistan conflicts. The cohort study was established in 2003 and has collected data over three previous phases including Phase 1 (2004-2006), Phase 2 (2007-2009) and Phase 3 (2014-2016).

**Methods and analysis:** Participants are eligible to take part if they completed the King’s Centre for Military Health Research (KCMHR) Health and Wellbeing Cohort Study at Phase 3 (2014-2016) and consented to be recontacted. Participants meeting these criteria will be recruited through email, post, and text message to complete an online or paper questionnaire. The study provides a fourth phase of quantitative longitudinal data on this cohort. Data are being collected between January 2022 and September 2023. Health and wellbeing measures used in Phase 4 include measures used in previous phases that assess common mental disorders (CMD), post-traumatic stress disorder (PTSD) and alcohol misuse. Other areas of interest assess multiple symptom illness, employment, help-seeking, and family relationships. New topics include the impact of the British withdrawal from Afghanistan in 2021, Complex-PTSD (C-PTSD), illicit drug use, gambling, and loneliness. The main analyses will compare mental health status according to deployment experiences and serving status (serving or ex-service) reporting prevalences with 95% Confidence Intervals (CI), and Odds Ratios (ORs) with 95% CI. Analyses will describe the effect size between groups deployed to Iraq and/or Afghanistan or not deployed, and those who are currently in service versus ex-service personnel respectively. Multivariable logistic and multiple linear regression analyses will be conducted to assess various health and wellbeing outcomes and associations with risk and protective factors, adjusting for potential confounders.

**Ethics and dissemination:** Ethical approval has been granted by the Ministry of Defence Research Ethics Committee (Ref: 2061/MODREC/21). Participants are provided with information and agree to a series of consent statements before taking part. Data are kept on secure servers and in locked cabinets/offices, with access to personally identifiable information limited. Findings will be disseminated to UK Armed Forces stakeholders and international research institutions through stakeholder meetings, project reports and scientific publications.

**Strengths and limitations of this study:**

- Strengths of this study include the original cohort recruitment from a random, representative sample of UK service personnel. Strengths also include recruitment from a cohort where underlying characteristics are known and longitudinal data are held on their health and wellbeing. The study has maintained validated and harmonised health and wellbeing measures across phases, whilst including new areas relevant to the cohort’s current experiences.
- This study will provide continued longitudinal data on this Armed Forces cohort.
- Study limitations include recruitment from a specific cohort; hence the study cannot comment on older era cohorts or those who joined the Armed Forces more recently.

## Introduction

### Context

The long-term health and wellbeing of serving (Regulars and Reservists) and ex-service personnel (veterans) is a key concern for the Government of the United Kingdom (UK), Ministry of Defence (MOD) and the Office for Veterans’ Affairs (OVA), and is a main tenet of their current strategies (1, 2). A focus on health and wellbeing throughout the life cycle of service in the Armed Forces helps support operational effectiveness and successful transitions to civilian life (3, 4). In addition, this emphasis on health and wellbeing fulfils commitments made in the Armed Forces Covenant (5) which aims to ensure that the UK Armed Forces community is not disadvantaged compared to the general population because of their military service. In 2022, a legal duty was placed on statutory bodies (government departments, agencies and public bodies) to give ‘due regard’ to the needs of the UK Armed Forces community (6). Hence, understanding the long-term health needs of current service and ex-service personnel is necessary to identify trends in health outcomes and inform prevention, early detection, and intervention strategies for this group. This evidence can also direct statutory and voluntary sector resources to members of the Armed Forces community who may need support.

In 2003, the King’s Centre for Military Health Research (KCMHR) Health and Wellbeing Cohort Study was set up to assess the possible consequences of deployment to the Iraq conflict (Operation TELIC) (7). Participants of the KCMHR Health and Wellbeing Cohort Study have been followed up over three main cohort timepoints with data collected between 2004-2006 (Phase 1), 2007-2009 (Phase 2) and 2014-2016 (Phase 3) (7-9). The cohort has also been utilised to collect extra waves of data such as data related to help-seeking, mental health of military fathers and their children, and the impact of the COVID-19 pandemic on the veteran community (10-13). Within the main waves of cohort data collection, during Phase 1, KCMHR surveyed a random sample of Regular and Reserve personnel who were deployed on Operation TELIC 1 and, for comparison, a random sample of trained but non-deployed personnel. At Phase 2, KCMHR followed up the original cohort, and in addition, included a random sample of Regular and Reservist personnel who had deployed to Afghanistan (Operation HERRICK), and a replenishment sample of personnel who had joined service since the cohort was first recruited in 2004. The replenishment sample was to ensure the cohort was broadly representative of the UK Armed Forces at the time of sampling. At Phase 3, all previous cohort participants were followed up and a further replenishment sample of personnel who joined the trained strength on or after 1 August 2009 was added to again ensure a broadly representative sample of the UK Armed Forces. By Phase 3 of the study, 40% of the sample had left service and transitioned to civilian life.

This study is a fourth wave of data collection (Phase 4 - 2022-2023) which includes both serving and ex-serving personnel (Regular and Reserves) participants who took part in Phase 3. In the UK and within this study, ex-service personnel/veterans are defined as anyone who has served for at least one day in His Majesty’s Armed Forces (Regular or Reservist) (14). In Phase 4 there is no replenishment sample of current Armed Forces, as the aim is to transition to a legacy cohort, broadly representative of those who served during the Iraq and Afghanistan era of conflicts and who share specific service, deployment, and combat experiences.

### Current knowledge

Whilst the majority of those who serve in the UK Armed Forces do not experience negative health outcomes from service, a significant minority may be at a higher risk of mental health problems and alcohol misuse compared to the UK general population (9). Risk factors for negative health outcomes include pre-service (childhood) vulnerabilities (15), exposures during service such as combat experiences (16), a military culture of alcohol use (17), and difficulties experienced during and after transition to civilian life, such as a poor integration into civilian life and limited social support (18). Service personnel and veterans may also experience increased barriers to seeking help for mental health conditions compared to the general population which affects access and engagement with healthcare services (11).

Over the three phases of the KCMHR cohort study, common mental disorders (CMD) were the most prevalent outcome in Regular service and ex-service personnel ranging between 20-22%, followed by alcohol misuse reported at 10-15% and probable post-traumatic stress disorder (PTSD) at 4-6% prevalence (7-9). CMDs have remained stable over the three phases, alcohol misuse has steadily declined, and probable PTSD increased at Phase 3 from 4% to 6%. Broadly, the prevalence of CMD and probable PTSD at Phase 3 were similar to general population estimates, with alcohol misuse rates approximately two to three times higher than equivalent general population estimates (9, 19). However, the prevalence of negative mental health outcomes was not uniform across groups, where ex-service personnel (compared to service personnel) who deployed in a combat role, reported higher levels of CMD, probable PTSD and alcohol misuse compared to equivalent general population estimates (9, 19, 20).

In Phases 1 and 2, deployed Reservists reported higher levels of probable PTSD compared to deployed Regulars (21), however this difference did not persist at Phase 3 with probable PTSD reported at 7% in both Reservists and Regulars. Deployed Reservists have consistently reported higher rates of CMD, PTSD and alcohol misuse compared to non-deployed Reservists (9).

Phase 4 of the study will be able to address questions regarding longer-term impacts of service, deployment and transition for this cohort that served during the Iraq and Afghanistan era of conflicts. The current phase will also provide new evidence in under-researched areas such as complex PTSD (C-PTSD) (22), gambling (23), illicit drug use (24), mild cognitive impairment (25), and will be able to assess impacts of events such as the withdrawal of troops from Afghanistan (26) and the COVID-19 pandemic (12, 27). The study will provide a robust evidence base for the UK government and stakeholders, such as Armed Forces charities, to help shape policy and support services. This study will be able to compare outcomes with previous phases and the general population, as well as with Armed Forces populations from allied countries.

### Research aims

The primary objective of Phase 4 of this longitudinal cohort study is to continue to describe the health and wellbeing of UK service and ex-service personnel, Regulars and Reservists, who served during the time of operations in Iraq (Operation TELIC) and Afghanistan (Operation HERRICK) and to describe the course of health outcomes over the Phases 1-4. Specific primary aims include being able to compare health outcomes between those who deployed to Iraq and/or Afghanistan and those that did not deploy, and between serving and ex-serving personnel. Other objectives are the assessment of a wider range of outcomes relevant to UK service and ex-service personnel. The study will assess:

- Post-service mental health, lifestyle behaviours, social support, and social exclusion.
- The predictors and associations of separation from service for both Regulars and Reservists.
- The prevalence of help-seeking for physical, mental health and alcohol problems and types of healthcare sources utilised.
- The prevalence and factors associated with problem gambling.
- The prevalence and factors associated with illicit drug use.
- Marital/relationship satisfaction and the effect of military service on relationships.
- The long-term health and social impact of alcohol misuse.
- The prevalence and factors associated with self-harm and suicidal ideation before, during, and after service.
- The prevalence and factors associated with Mild Traumatic Brain Injury (mTBI) and post-concussion symptoms (PCS).
- The prevalence and factors associated with anger, aggressive behaviour, and interpersonal violence.
- The prevalence and factors associated with mild cognitive decline.
- The impact of the British withdrawal from the NATO mission in Afghanistan on the health and wellbeing of service and ex-service personnel.

## Method and analysis

### Study protocol

#### Sample population and eligibility

Participants will be recruited from those who responded at Phase 3 of the cohort study and who consented to future contact (N = 7608).

#### Data collection method and timeline

Data will be collected via an online or paper questionnaire. The online questionnaire will be accessed through Qualtrics software (28) and will take approximately 40-45 minutes to complete. Participants invited to take part provided contact details during previous phases of the study. The MOD will provide updated contact details for participants who were in-service at the time of Phase 3. This data linkage will be carried out under a data usage agreement between the MOD and KCMHR which adheres to data protection legislation.

Data collection will be carried out over a period of approximately 20-22 months, starting January 2022 and anticipated to close in September 2023. Participants will be given as much time as they like to decide if they wish to participate up until the end of data collection.

#### Initial contact

In the first instance, participants who have provided a personal email address at a previous phase will be invited to take part in the online survey by email. The email invite emphasises that participation is voluntary, confidential, and will include a personalised questionnaire link, a link to the Participant Information Sheet (PIS), and a link to the study website. Where we do not hold a personal email address, participants will be invited to take part by post. An initial invite letter will be sent detailing how to take part in the study online.

Where email or postal addresses are found to be invalid, we will send invitations to alternative contact details that we hold, and we may also send a text message where possible to participants to invite them to take part and provide details of how to find out more about study.

#### Reminder invitations

Up to two reminders will be sent to those who have not completed the survey and have not refused participation. For those who have been contacted by email, a first reminder will be sent approximately two weeks after the initial invitation, and a second reminder approximately two weeks later. For those who have been invited to take part by post, we will send a postal pack including a personal link to take part in the online survey and a paper questionnaire, a reply-paid return envelope and a PIS. This will be mailed to either the most recent address held on our database, or any updated addresses provided by the MOD.

#### Follow up and tracing of non-responders

Following the reminder invitations, several methods will be used to follow up and trace non-responders. We will attempt to contact non-responders by telephone to check whether participants have received the study invitation and to answer any queries they may have about the study. Those who wish to take part will be sent a new invitation by email, text, or post as preferred. If we are unable to reach participants by telephone or telephone numbers are no longer valid, we will check Directory Enquiries for updated numbers.

#### ‘Final Chance’ contact

Approximately one month before the end of data collection, all non-responders will be sent a ‘final chance’ email, text or letter detailing that the study data collection will close and describe how they can take part in the survey online or to request a paper questionnaire.

#### Other communication activities

Information about the study will be shared on the KCMHR website (KCMHR.org), through blogs, through social media posts on the KCMHR Twitter profile and through Armed Forces stakeholders’ newsletters and websites. Newsletters will be shared with the cohort members during data collection, informing participants of progress and providing further information about the study.

#### Study materials

The study will provide a website giving more information on the study and links to the PIS, Signposting Booklet, and privacy notice. The PIS provides detailed study information, answers to frequently asked questions and contact details for the research team. The options regarding refusal to take part and withdrawal from the study are explained in the PIS. The Signposting Booklet offers information about relevant support services. A link to the Signposting Booklet will be provided in the online survey and details will also be included in the paper questionnaire pack. A risk protocol is in place for participants that get in touch with the study team who are distressed and in need of sign posting or immediate crisis help.

### Measures

As we are seeking to assess change, the principal outcome measures are the same screening measures used in previous phases (numbered 1 to 10). New measures at Phase 4, reflecting current topics of interest are numbered 11-18.

1. 12-item General Health Questionnaire (GHQ-12) as a measure of general (non-psychotic) psychiatric morbidity. Cut off indicating the presence of a CMD such as depression or anxiety ≥4 (29).
2. 17-item PTSD Checklist (PCL-C) to measure probable PTSD. Cut off for probable PTSD ≥50 (30).
3. 20-item PCL-5 to measure probable PTSD. Cut off for probable PTSD ≥38 (31).
4. 10-item Alcohol Use Disorder Identification Test (AUDIT) to measure alcohol consumption and misuse. Cut off for alcohol misuse ≥16 (32).
5. 15-item Patient Health Questionnaire (PHQ-15) to measure multi-symptom illness. Cut off for multi-symptom illness ≥10 (33).
6. Two items from the Brief Traumatic Brain Injury Screen (BTBIS) as a measure of mild traumatic brain injury (mTBI) (34).
7. Domestic abuse questions from Adult Psychiatric Morbidity Survey 2014 (APMS) (19).
8. Three items selected from the Short Form (SF36) Quality of Life measure (35).
9. 5-item Dimensions of Anger Reactions (DAR-5). Cut off ≥12 to denote anger difficulties (36).
10. 4-item measure of aggressive behaviour used by the Walter Reed Army Institute of Research (37).
11. 4-item Drug Use Disorders Identification Test (DUDIT). Cut off for drug related problems >5 (38).
12. Three suicidal ideation/self-harm items taken from the Clinical Interview Schedule-Revised (CIS-R)12 and used in the Adult Psychiatric Morbidity Survey (39).
13. Problem Gambling Severity Index (PGSI). Cut off for problem gambling ≥8 (40).
14. 4-item Dyadic Adjustment Scale (DAS-4) of marital/relationship quality (41).
15. 3-item UCLA Loneliness Scale. Cut off for loneliness ≥6 (42).
16. Oslo Social Support Scale (OSSS-3) (43), scoring as per Bøen, Dalgard (44).
17. The International Trauma Questionnaire (ITQ) to measure C-PTSD, scoring as per Cloitre, Shevlin (45) .
18. 16-Item Informant Questionnaire on Cognitive Decline (IQCODE Short Form). Scores summed and divided by 16, cut off ≥3.31 (46).

From 2013, the definition of PTSD changed from DSM-IV to DSM-5. A new PCL-5 measure was introduced in our cohort to reflect the changes. To continue to compare the level of probable PTSD across phases but to also be able to report current probable PTSD using the new definition, a blended PCL measure will be included that allows the creation of both a PCL-C and a PCL-5 measure.

Other key topics will include questions regarding social and military demographic characteristics, employment, finances, accommodation and living arrangements, experience of Reserve service, help-seeking for health problems, impact of the Afghanistan withdrawal, and COVID-19.

### Power calculations

At Phase 3, the response rate among those who had participated in previous phases was 58%. Among those who were new to the study (replenishment group), the response rate was lower. However, at Phase 4 we will only include those who participated at Phase 3, i.e., there will be no new participants. We therefore anticipate a response rate of at least 50-60% at Phase 4. With 7608 available for recontact, a response rate of 60% would result in 4565 participants at Phase 4.

The statistical power and sample sizes for all phases have been based on the prevalence of probable PSTD which, within this study, is the outcome of interest with the lowest prevalence (7-9). The planned study design will allow us to conduct two main analyses: a) cross-sectional and b) longitudinal.

#### Cross-sectional analyses

A primary aim is to continue to monitor the mental health of personnel who have deployed on operations to Iraq and Afghanistan compared to personnel who did not deploy on those operations. Assuming 4565 responses at Phase 4 and assuming 62% will have deployed to Iraq or Afghanistan (as at Phase 3): deployed (n = 2830), non-deployed (n = 1735). Assuming a prevalence of PTSD of 5.0% in the non-deployed group (prevalence at Phase 3 (9)), we would have more than 90% power to detect a 2.5% difference in the deployed group compared to the non-deployed group. This calculation is based on the difference between two proportions with 95% confidence (two−sided) and based on unequal sample sizes.

A second primary aim is to compare the prevalence of PTSD in serving and ex-service personnel. Assuming 4565 responses at Phase 4 and assuming 70% will have left service: ex-service (n = 3196), serving (n = 1369). Assuming a prevalence of PTSD of 4.6% in the serving group (prevalence at Phase 3), we would have more than 90% power to detect a 2.5% difference in the ex-service group compared to the serving group. This calculation is based on the difference between two proportions with 95% confidence (two−sided) and based on unequal sample sizes.

#### Longitudinal analyses

The aim is to detect a change in the prevalence of PTSD at Phase 4 compared to previous phases in the whole sample. Assuming a prevalence for probable PTSD of 6.2% (prevalence in whole sample at Phase 3), a sample size of 4565 pairs would have over 90% power to detect an increase prevalence to 7.7% (difference in proportions of.0.015) with an alpha of 0.05 (two-sided). This is assuming that two thirds of the original 6.2% will recover (47). This calculation is based on a McNemar’s test of equality of paired proportions with 95% confidence (two-sided).

### Statistical methods

Statistical analysis will be carried out using the statistical packages Stata, SPSS and Mplus. The main analyses will compare mental health status according to deployment experiences and serving status, reporting prevalences with 95% CI, and Odds Ratios (ORs) with 95% CI, to describe the effect size between groups deployed to Iraq and/or Afghanistan or not deployed, and those who are current service personnel versus those who are ex-service personnel respectively. We will use paired t−tests for continuous outcomes, and McNemar’s tests for dichotomous outcomes, to compare prevalence in outcomes between phases. Multivariable logistic and multiple linear regression analyses will be conducted to assess mental health outcomes and associations with risk and protective factors. Regression analyses will control for a priori confounders identified relevant to the specific research question e.g., age, sex, rank, service branch, enlistment status (Regular/Reserve).

Further analyses will determine risk factors for new cases of key outcomes based upon responses at previous phases, as well as factors associated with improvement of symptoms, using multiple logistic regression and multinomial logistic regression. Latent class growth analysis (LCGA) and growth mixture modelling (GMM) will be used to study trajectories of, for example, PTSD and alcohol use. Structural equation modelling will be used to assess mediators and indirect effects.

The response rate will be calculated as per previous phases (7-9) (number responding divided by number in sample at Phase 4, multiplied by 100). We will report an adjusted response rate calculated as the number responding divided by the number in sample at phase 4, minus number of deaths and those who are uncontactable due to incorrect contact details, multiplied by 100. Response weights will be calculated as the inverse probability of responding once sampled (at the appropriate phase) driven by factors shown empirically to predict response. Combined sample and response survey weights will be used to account for differential response rates and sampling fractions in all analyses. All screening measures will be scored according to published instructions. Missing items on measures will be handled according to the protocol used in the previous phases, i.e., for example on GHQ-12, PCL-17, PCL-5 and AUDIT, the lowest value is imputed if fewer than four items are missing on the measure.

### Patient and public involvement

Veterans from the Veterans Research Advisory Group tested the design and flow of the questionnaire and offered advice on outcome measures. The Advisory Group were specifically consulted on questions and items assessing the impact of the British withdrawal from the NATO mission in Afghanistan. Results of the study will be disseminated to participants through a newsletter, our social media outlets, and through stakeholders that represent Armed Forces communities.

### Data storage and security

#### Database

Personal information which identifies individuals (such as date of birth, service number) will be stored in an encrypted database on a restricted server, accessible only within KCMHR offices, and only by specified, authorised, members of the team. Access to the server is via an allocated login and password, which is unique for each researcher.

#### Pseudonymisation of questionnaire data

Individuals were assigned a unique ID by the MOD at the point of sampling. A second unique ID number has been generated by KCMHR to identify records in datasets for analysis allowing survey data to be held pseudonymously. Questionnaire data will be stored separately to the identifiable data in both electronic and paper format. Hard copy versions of the questionnaire will include a ‘contact sheet’ enabling participants to provide preferred contact details and consent information. The contact sheet will be removed and stored separately from the questionnaire; both will be labelled with the participant ID number.

#### Data entry of paper questionnaires

The hard copy questionnaire data and contact form will be entered in-house by the research team using Qualtrics. The paper questionnaire will be checked for inaccuracies and unclear responses which will be flagged and then reviewed by the Project Manager.

#### Data management and oversight

All research staff are provided with training regarding the General Data Protection Regulation (GDPR), King’s College London (KCL) standards for handling data and Medical Research Council (MRC) data protection training (48).

Data required for analysis will be extracted from the KCMHR secure system in pseudonymised form and stored on KCL OneDrive. We will use a valid Data Protection and Security Toolkit from KCMHR at KCL and will follow this Toolkit during rollout. Pseudo-anonymous datasets may also be shared with other Institutions under Data Sharing Agreements. This will be stated in the Participant Information Sheet and study privacy notice. Data will be kept by KCMHR for twenty years in line with MRC guidelines and we may use the pseudonymous data in future research.

## Ethics and Dissemination

Full ethical approval has been granted by the UK Ministry of Defence Research Ethics Committee (Ref: 2061/MODREC/21).

### Financial incentives

As in previous phases, participants who complete a questionnaire will be given the opportunity to be entered into a prize draw to win an amount between £25 and £1000, either for themselves or a charity of their choice. The amount and number of each prize available will be clearly stated in the participant information. The prizes available will be 1 x £1000, 2 x £500, 5 x £100, 5 x £50, 10 x £25. Entry into the prize draw is voluntary.

## Dissemination

Results will be written up in academic papers and submitted for peer reviewed publication. No identifying information will be presented in any reports or publications. Aggregated data will be made available in a final report to the funder and in future academic publications, but consideration will be given to cells with small numbers and may be supressed. A final study report will be presented to the OVA, and academic publications will be disseminated through KCL communications channels in social media, press briefings, and academic conferences.

## Conclusions

The Health and Wellbeing Study of serving and ex-serving UK Armed Forces personnel is a longitudinal cohort study investigating the impact of service in the Armed Forces on those who served during the time of the recent Iraq and Afghanistan conflicts. The study has three main previous data collection phases (2004-2006, 2007-2009 and 2014-16). This current (fourth) phase of data collection (2022-2023) will recruit from participants who took part in Phase 3 of the study. The Phase 4 sample includes service personnel (Regulars and Reservists) and ex-service personnel. The study strengths include recruitment from a population where underlying characteristics are known, providing longitudinal data on health and wellbeing, and the use of validated measures for mental health and wellbeing outcomes. Study limitations include recruitment from a specific cohort that served during the Iraq and Afghanistan conflicts.

## Data Availability

Data are available upon reasonable request. Data will be processed in accordance with the General Data Protection Regulation (GDPR) and the Data Protection Act 2018. We will not make any record-level data publicly accessible because we need to protect the confidentiality and security of the individual cohort members. You are welcome to contact us with proposals for collaborative research, which the investigators will consider on a case-by-case basis, and which will only occur as part of a legal collaborative agreement and after the collaborator has put in place the relevant research ethics, data protection and data access approvals.

## Authors’ Contributions

MLS, MJ, RL, LH, HB, AS, DL, NG, DM, DMacM, SW, SAMS, NF were involved in the original concept and design of the study and questionnaire. NTF, SAMS and SW have overseen the conduct of all aspects of the study. RL and DL led on online survey design, format, and flow. MJ, LH and MLS led on the ethics submission with substantial contributions from RL, HB, AS, DL, NG, DM, SW, SAMS and NTF. MJ and LH led on the design of participant materials including the participant invite and information sheet with input from all authors. MJ and MLS led on the data analysis plan with input from HB, SW, SAMS and NTF. MLS led the writing of the protocol paper, with drafting and revision input from all named authors. All named authors have all approved the final version of this paper and accept accountability for all aspects of the work.

## Acknowledgments

Thank you to all our KCMHR colleagues and the veterans who helped to edit and shape the survey questionnaire.

## Ethics approval and consent to participate

Full ethical approval has been granted by the UK Ministry of Defence Research Ethics Committee (Ref: 2061/MODREC/21) and all participants will provide informed consent before participating. Participants are provided with information and agree/disagree to a series of online or written consent statements before completion of the study questionnaire. Participants are also provided with information on how to withdraw from the study if they wish to after participation up until the end of data collection. Please note CONSORT guidelines are not applicable.

## Availability of data and materials

For the purposes of open access, the author has applied a Creative Commons Attribution (CC BY) licence to any Accepted Author Manuscript version arising from this submission.

